# Distinct COVID-19 vaccine combinations result in divergent immune responses

**DOI:** 10.1101/2023.08.25.23294606

**Authors:** Luca M. Zaeck, Ngoc H. Tan, Wim J.R. Rietdijk, Daryl Geers, Roos S.G. Sablerolles, Susanne Bogers, Laura L.A. van Dijk, Lennert Gommers, Leanne P.M. van Leeuwen, Sharona Rugebregt, Abraham Goorhuis, Douwe F. Postma, Leo G. Visser, Virgil A.S.H. Dalm, Melvin Lafeber, Neeltje A. Kootstra, Anke L.W. Huckriede, Bart L. Haagmans, Debbie van Baarle, Marion P.G. Koopmans, P. Hugo M. van der Kuy, Corine H. GeurtsvanKessel, Rory D. de Vries, SWITCH-ON Research Group

**Author notes:** Contributed equally (Luca M. Zaeck, Ngoc H. Tan). Contributed equally (P. Hugo M. van der Kuy, Corine H. GeurtsvanKessel, Rory D. de Vries). List of investigators from the SWITCH-ON research group is in **Supplementary Table S1**. Corresponding author: Corine H. GeurtsvanKessel, department of Viroscience, Erasmus Medical Center, Rotterdam, 3015 GD, Netherlands.

## Abstract

Waning antibody responses after COVID-19 vaccination combined with the emergence of the SARS-CoV-2 Omicron lineage led to reduced vaccine effectiveness. As a countermeasure, bivalent mRNA-based booster vaccines encoding the ancestral spike protein in combination with that of Omicron BA.1 or BA.5 were introduced. Since then, BA.2-descendent lineages have become dominant, such as XBB.1.5 or BA.2.86. Here, we assessed how different COVID-19 priming regimens affect the immunogenicity of the recently used bivalent booster vaccinations and breakthrough infections. BA.1 and BA.5 bivalent vaccines boosted neutralizing antibodies and T-cells up to 3 months after boost; however, cross-neutralization of XBB.1.5 was poor. Interestingly, different combinations of prime-boost regimens induced divergent responses: participants primed with Ad26.COV2.S developed lower binding antibody levels after bivalent boost while neutralization and T-cell responses were similar to mRNA-based primed participants. In contrast, the breadth of neutralization was higher in mRNA-primed and bivalent BA.5 boosted participants. Combined, we highlight important ‘lessons learned’ from the employed COVID-19 vaccination strategies. Our data further support the use of monovalent vaccines based on circulating strains when vaccinating risk groups, as recently recommended by the WHO. We emphasize the importance of the continuous assessment of immune responses targeting circulating variants to guide future COVID-19 vaccination policies.

## Introduction

Vaccination against coronavirus disease-2019 (COVID-19) provides protection against infection, hospitalization, and mortality^1,2^. However, ongoing waning of severe acute respiratory syndrome coronavirus-2 (SARS-CoV-2)-specific immune responses and the continuous evolution of antigenically distinct variants result in an overall reduction of vaccine effectiveness^3^. The currently circulating Omicron BA.2-descendent variants such as XBB.1.5 are the most immune evasive to date^4–6^. This leads to an ongoing arms race: adapted vaccines are required to retain effective protection on a population level, especially in vulnerable at-risk patients, in the face of new emerging variants. As a countermeasure, mRNA-based bivalent vaccines incorporating an Omicron BA.1 or BA.5 spike (S) protein in combination with the ancestral S were introduced in 2022^7,8^.

While mRNA-based vaccines, including BNT162b2 and mRNA-1273, were initially shown to have higher vaccine efficacy over adenovirus-vectored vaccines (Ad26.COV2.S and ChAdOx1-S) in a primary vaccination series^3,9^, it is not known whether the different original priming regimens have a long-lasting imprinting effect on the magnitude, durability, or breadth of the SARS-CoV-2-specific immune response^10^. Heterologous COVID-19 vaccination with different vaccine platforms but the same S antigen was demonstrated to be at least non-inferior regarding immunogenicity when compared to homologous priming with either mRNA-based or adenovirus-based vaccines alone^11–13^. Shaping of the immune response as a consequence of exposure to different S antigens was mostly studied in the context of hybrid immunity, a combination of vaccination and infection. These studies showed evidence for serological imprinting to the ancestral S protein, but also the induction of variant-specific immune responses^14–16^.

The SWITCH-ON trial^17,18^ aimed to evaluate the mRNA-based bivalent BA.1 and BA.5 booster vaccines developed by Pfizer (BNT162b2) or Moderna (mRNA-1273.214 and mRNA-1273.222) against the background of different priming regimens (mRNA-based or adeno-based), by addressing three crucial questions: (1) How immunogenic are Omicron BA.1 or BA.5 bivalent booster vaccines? (2) Do BA.1 or BA.5 bivalent booster vaccines differ in the induction of broad neutralizing antibody responses, including adequate neutralization of XBB-descendent variants? (3) How do immune responses among different original priming vaccination regimens evolve over time and what can we learn for the future?

## Results

### Study design and baseline characteristics

A total of 434 healthcare workers (HCW) were included in the SWITCH-ON trial after screening of 592 potential participants (**Figure 1**, baseline characteristics in **Supplementary Tables S2** and **S3**). HCW received either Ad26.COV2.S or an mRNA-based (mRNA-1273 or BNT162b2) priming vaccination regimen, followed by at least one mRNA-based booster vaccination before inclusion in this study. The SWITCH-ON trial comprised two groups to which the participants were randomly assigned: (1) a direct boost group (DB) (n=219, accounting for dropouts) or (2) a postponed boost (PPB) group (n=183, accounting for dropouts). Participants in the DB group were vaccinated in October 2022 with an Omicron BA.1 bivalent vaccine (BNT162b2 Omicron BA.1 or mRNA-1273.214); participants in the PPB group were vaccinated in December 2022 with an Omicron BA.5 bivalent vaccine (BNT162b2 Omicron BA.5 or mRNA-1273.222). Samples were collected before bivalent vaccination, at 7 and 28 days post-vaccination, and at approximately 3 months post-vaccination (**Supplementary Figure S1**). No formal statistical tests were performed to test for differences within or between groups as we deviated from the original protocol in terms of pre-specified outcomes and a lower than anticipated sample size^18^.

**Figure 1.**
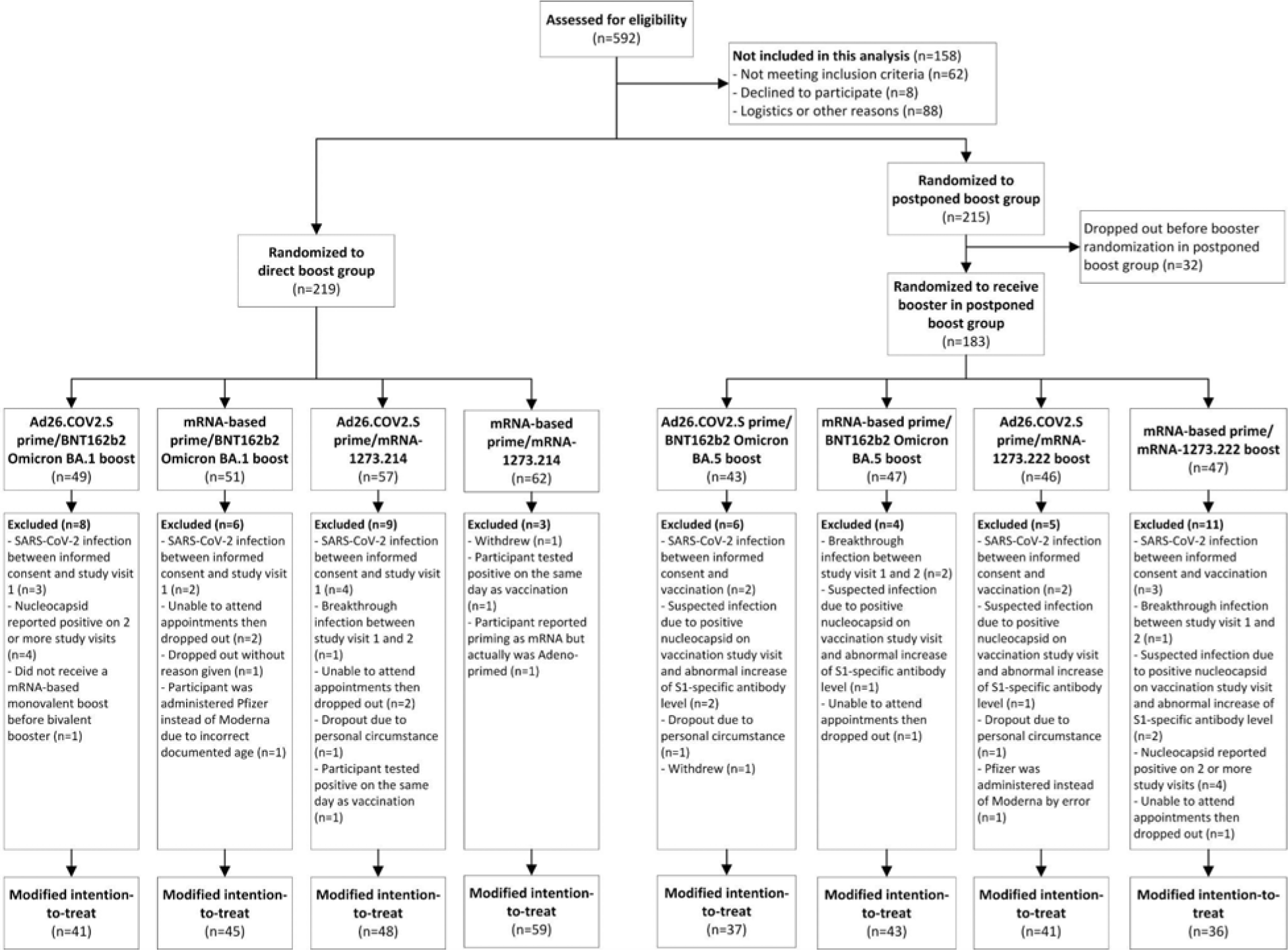
SWITCH-ON trial enrollment. A total of 592 healthcare workers (HCW) were screened for eligibility, of whom 434 were included and randomized 1:1 to the direct boost (n = 219) and the postponed boost (n = 215) group. Following dropouts, a total of 183 HCW received an Omicron BA.5 bivalent vaccine in the postponed group.

### Bivalent COVID-19 vaccines induce antibody and T-cell responses

The immunogenicity of Omicron BA.1 bivalent vaccines up to 28 days post-vaccination in the SWITCH-ON trial was reported previously^17^. Both S-specific IgG binding and neutralizing antibodies targeting ancestral SARS-CoV-2 increased within the first 28 days, with most of the increase occurring between day 0 and 7 (**Figure 2a,b**). S-specific T-cell responses increased rapidly in the first 7 days post-vaccination and subsequently waned (**Figure 2c**). At 3 months post-vaccination, all of the measured immune parameters had decreased in comparison to the previous study visit. Whereas antibodies did not yet wane to baseline levels, T-cell responses returned close to baseline. The magnitude and kinetics of antibody and T-cell responses induced by Omicron BA.5 bivalent booster vaccination were comparable to Omicron BA.1 bivalent boost, again with most of the increase occurring within the first 7 days (**Figure 2d-f**). Overall, a comparable boost of (neutralizing) antibody and T-cell responses against ancestral SARS-CoV-2 was observed after either Omicron BA.1 or BA.5 bivalent boost, independent of the timing of vaccine administration.

**Figure 2.**
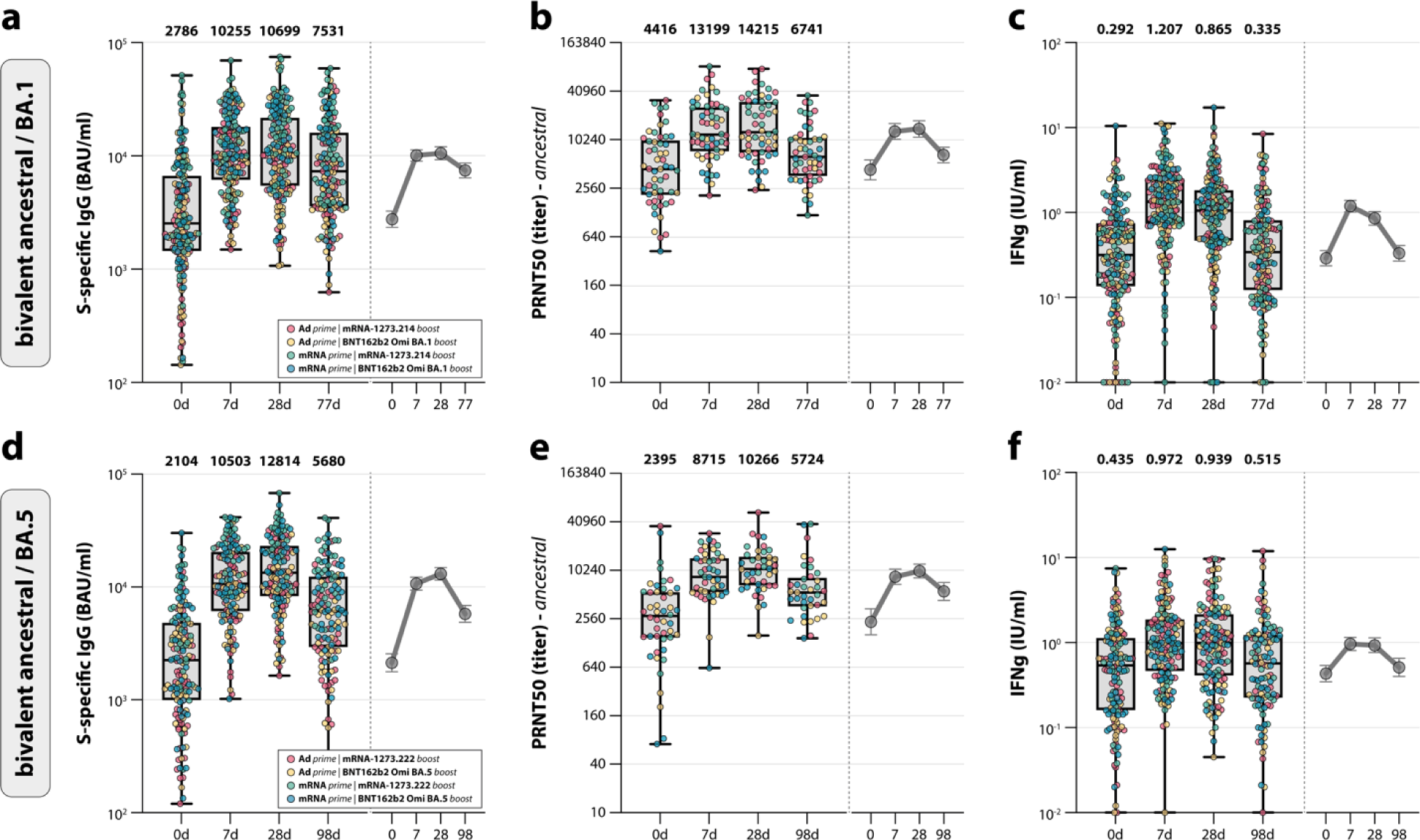
Antibody and T-cell responses after bivalent booster vaccination. **a-f**, Detection of (ancestral) spike (S)-specific binding IgG antibodies (**a,d**), ancestral SARS-CoV-2 neutralizing antibodies (**b,e**), and T-cell responses measured by interferon-gamma (IFN-γ) release assay (IGRA) (**c,f**) after Omicron BA.1 (**a-c**) or BA.5 (**d-f**) bivalent booster vaccination at baseline, and 7 days, 28 days, and 3 months post-boost. Colors indicate the specific prime-boost regimen (red = Ad26.COV2.S prime, mRNA-1273.214 or mRNA-1273.222 boost; yellow = Ad26.COV2.S prime, BNT162b2 Omicron BA.1 or BA.5 boost; green = mRNA-based prime, mRNA-1273.214 or mRNA-1273.222 boost; blue = mRNA-based prime, BNT162b2 Omicron BA.1 or BA.5 boost). Data are shown in box-and-whisker plots, with the horizontal lines indicating the median, the bounds of the boxes indicating the IQR, and the whiskers indicating the range. Bold numbers above the plots represent the respective geometric mean (titer) per timepoint. The line graphs next to each panel depict a time course of the respective geometric mean values with 95% confidence intervals.

### mRNA-based priming leads to higher binding antibody levels after bivalent boost

The two groups (DB, Omicron BA.1 bivalent boost; PPB, Omicron BA.5 bivalent boost) could each be subdivided into four subgroups, based on different priming and bivalent booster regimens: (1) Ad26.COV2.S prime and mRNA-1273.214 or mRNA-1273.222 boost, (2) Ad26.COV2.S prime and BNT162b2 Omicron BA.1 or BA.5 boost, (3) mRNA (mRNA-1273 or BNT162b2)-based prime and mRNA-1273.214 or mRNA-1273.222 boost, and (4) mRNA-based prime and BNT162b2 BA.1 or BA.5 boost (**Supplementary Figure S1**). Notably, Omicron BA.1- and BA.5-boosted participants who had previously received an mRNA-based priming vaccination regimen consistently had higher levels of S-specific binding antibodies than those who received an Ad26.COV2.S priming (**Figure 3a,b**, compare green and blue to red and yellow). This effect of the original priming was not observed for ancestral SARS-CoV-2 neutralizing antibodies or T-cell responses (**Supplementary Figure S2**). Of specific interest, bivalent booster vaccination with mRNA-1273.214 or mRNA-1273.222 resulted in a larger increase of binding and neutralizing antibodies than boosting with their BNT162b2 counterparts did, indicating that these vaccines are more immunogenic (**Figure 3** and **Supplementary Figure S3**). Both findings underline that different prime-boost regimens lead to divergent immune responses.

**Figure 3.**
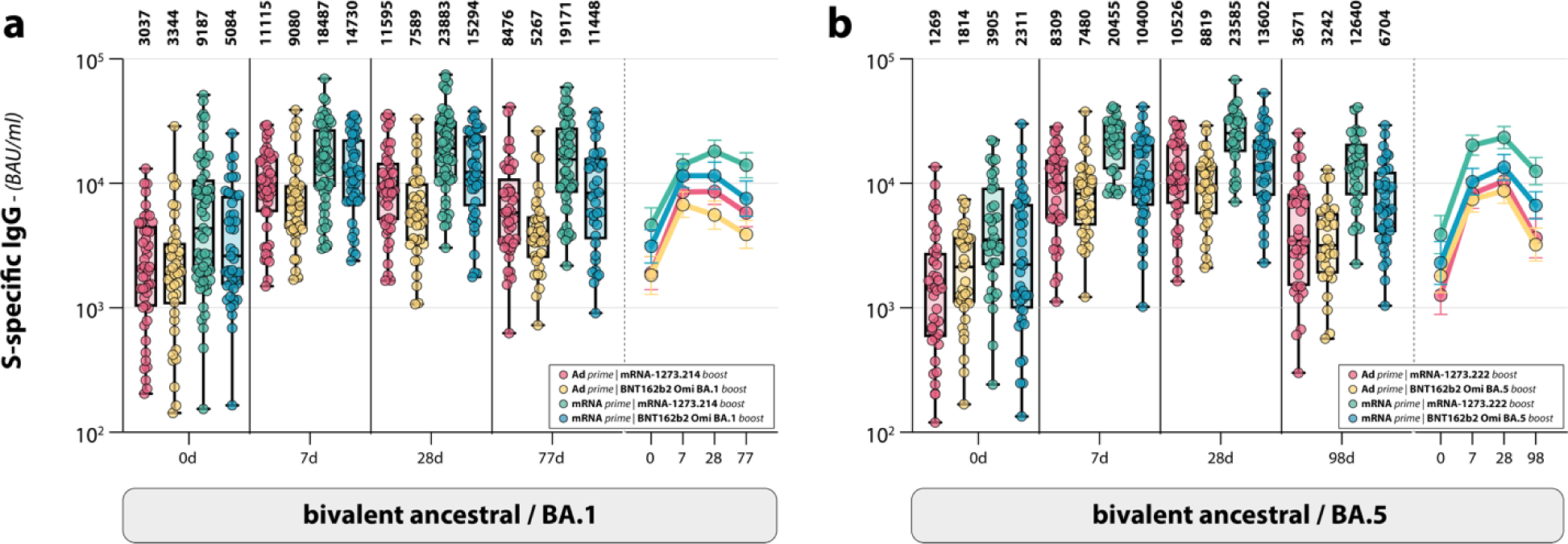
Antibody and T-cell responses after different original priming and bivalent booster vaccinations. **a,b**, Detection of S-specific binding IgG antibody levels in subgroups based on the different combinations original priming regimen after Omicron BA.1 (**a**) or BA.5 (**b**) bivalent booster vaccination at baseline, and 7 days, 28 days, and 3 months post-boost (red = Ad26.COV2.S prime, mRNA-1273.214 or mRNA-1273.222 boost; yellow = Ad26.COV2.S prime, BNT162b2 Omicron BA.1 or BA.5 boost; green = mRNA-based prime, mRNA-1273.214 or mRNA-1273.222 boost; blue = mRNA-based prime, BNT162b2 Omicron BA.1 or BA.5 boost). Data are shown in box-and-whisker plots, with the horizontal lines indicating the median, the bounds of the boxes indicating the IQR, and the whiskers indicating the range. Bold numbers above the plots represent the respective geometric mean (titer) per timepoint. The line graphs next to each panel depict a time course of the respective geometric mean values with 95% confidence intervals.

### mRNA-based prime followed by BA.5 bivalent boost leads to broad neutralization

Neutralizing antibodies against relevant Omicron variants BA.1 and BA.5 (encoded by the vaccines), and XBB.1.5 (circulating) were measured to assess the breadth of the neutralization response (**Figure 4a,b**). Comparable to ancestral SARS-CoV-2 neutralization, Omicron BA.1 and BA.5 neutralization were boosted by both the BA.1 and BA.5 bivalent booster vaccines; however, levels remained below those for ancestral SARS-CoV-2 neutralization at all timepoints. At 3 months post-vaccination, waning of neutralizing antibodies was observed. Remarkably, when correlating ancestral- and variant-specific neutralizing antibody titers (**Supplementary Figure S4**), it was clear that waning of Omicron BA.1 and BA.5 neutralizing antibodies occurred at a slower rate compared to ancestral SARS-CoV-2 neutralizing antibodies. This was true for both individuals boosted with the bivalent Omicron BA.1 (**Figure 4c,e**) or BA.5 vaccine (**Figure 4d,f**). The circulating Omicron XBB.1.5 was poorly cross-neutralized at 3 months after bivalent boost, irrespective of the different prime-boost regimens (**Figure 4a,b**). In participants boosted with the bivalent Omicron BA.5 vaccine, a preferential boost of Omicron BA.5 neutralization was observed. This was not the case for Omicron BA.1 neutralizing antibodies in participants boosted with the bivalent Omicron BA.1 vaccine (**Figure 4g**, compare orange with purple radar plot). When subdividing participants boosted with bivalent Omicron BA.5 in their respective prime-boost regimens, preferential boosting of Omicron BA.5 neutralization was restricted to participants who were primed with an mRNA-based vaccine (**Figure 4h**). Participants primed with Ad26.COV2.S retained a narrow neutralizing response, despite receiving the bivalent Omicron BA.5 booster.

**Figure 4.**
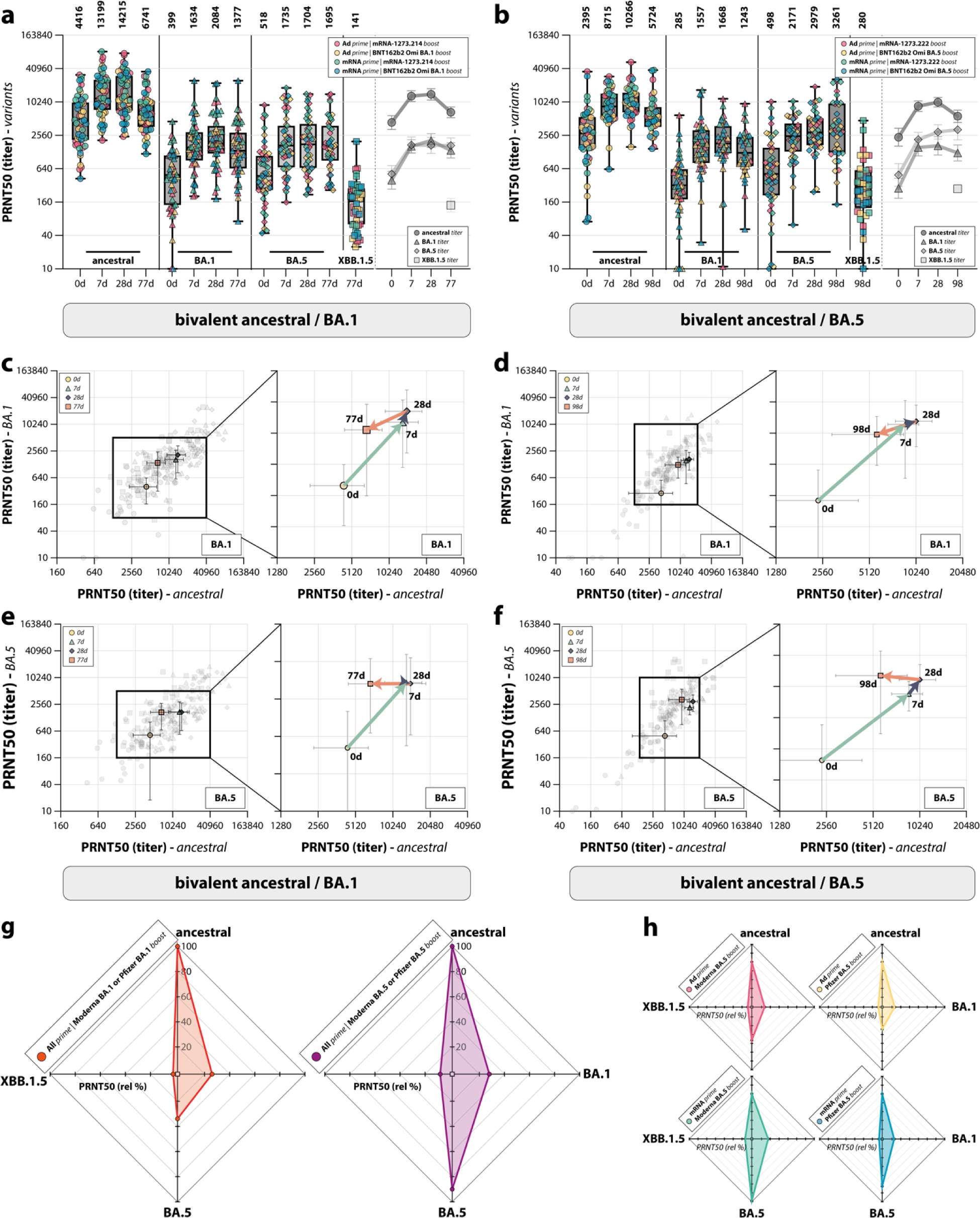
Breadth of the neutralizing antibody response after bivalent booster vaccination. **a,b**, Detection of neutralizing antibodies targeting ancestral SARS-CoV-2 and Omicron BA.1, BA.5, and XBB.1.5 variants after Omicron BA.1 (**a**) or BA.5 (**b**) bivalent booster vaccination at baseline, and 7 days, 28 days, and 3 months post-boost. Colors indicate the specific prime-boost regimen (red = Ad26.COV2.S prime, mRNA-1273.214 or mRNA-1273.222 boost; yellow = Ad26.COV2.S prime, BNT162b2 Omicron BA.1 or BA.5 boost; green = mRNA-based prime, mRNA-1273.214 or mRNA-1273.222 boost; blue = mRNA-based prime, BNT162b2 Omicron BA.1 or BA.5 boost). **c-f**, Correlation between PRNT50 titers against ancestral SARS-CoV-2 and the Omicron BA.1 (**c,d**) or BA.5 (**e,f**) variants over time after Omicron BA.1 (**c,e**) or BA.5 (**d,f**) vaccination at baseline, and 7 days, 28 days, and 3 months post-boost. Colored symbols indicate the specific timepoints (yellow = baseline [0 d]; teal = 7 d; purple = 28 d; orange = 77 d [**c,e**]/98 d [**d,f**]). The arrows connect the correlated geometric means (+ 95% CI) per timepoint and visualize the neutralization kinetics. **g,h**, Spiderweb plots depicting the variant-specific PRNT50 titers relative to ancestral SARS-CoV-2 neutralization (set to 100%), after vaccination with bivalent Omicron BA.1 (**g**) or BA.5 (**g,h**). Data in panels **a,b** are shown in box-and-whisker plots, with the horizontal lines indicating the median, the bounds of the boxes indicating the IQR, and the whiskers indicating the range. Bold numbers above the plots represent the respective geometric mean (titer) per timepoint. The line graphs next to each panel depict a time course of the respective geometric mean values with 95% confidence intervals.

### Breakthrough infections lead to comparatively low levels of immune responses

In the PPB group, which was included in September 2022 but scheduled to receive the bivalent Omicron BA.5 vaccine in December 2022, 13 test-confirmed infections were detected before administration of the booster dose (**Figure 5a**). These participants were subsequently excluded from the vaccine trajectory and analyzed separately as part of a natural infection-related sub-study. Breakthrough infection before bivalent vaccination boosted S-specific binding antibodies and T-cell responses. However, binding antibody levels 7 days (GMT 3,655 BAU/mL [95% CI 2,167-6,166]) and 28 days (GMT 5,025 BAU/mL [95% CI 3,264-7,737]) post-infection (**Figure 5b**) were considerably lower than compared to the same time interval post-vaccination (7d: GMT 10,503 BAU/mL [95% CI 9,218-11,969]; 28d: GMT 12,814 BAU/mL [95% CI 11,271-14,569], shown in **Figure 2d**). T-cell responses and Omicron neutralizing antibodies were comparable to post-vaccination responses, although T-cell responses returned to baseline faster compared to post-vaccination (**Figure 5c,d**). In addition, 59 breakthrough infections after administration of either bivalent Omicron BA.1 or BA.5 booster vaccination were detected through various methods (test-confirmed or detection of nucleocapsid-specific antibodies). Of these participants, samples collected prior to infection were included in the preceding immunogenicity analyses. Notably, breakthrough infection after Omicron BA.1 or BA.5 bivalent boost did not result in an additional increase of antibody or T-cell responses in comparison to the already vaccine-induced levels (**Supplementary Figure S5**). Overall, breakthrough infections before and after vaccination were comparatively poorly immunogenic compared to vaccine-induced immune responses.

**Figure 5.**
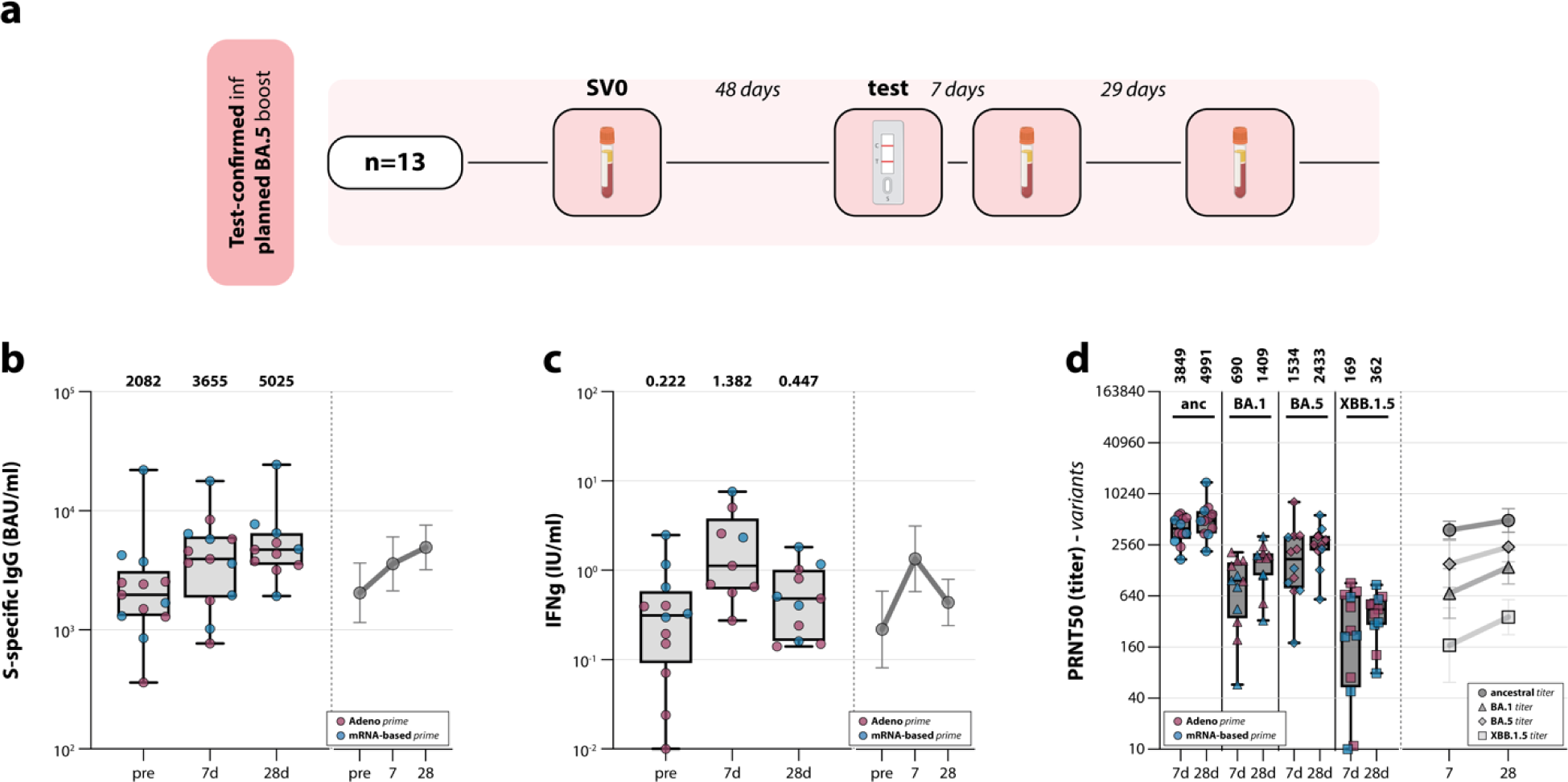
Breadth of the neutralizing antibody response after breakthrough infection. **a**, Sampling procedure for participants in the postponed boost group who had a breakthrough infection before their intended vaccination with the bivalent Omicron BA.5 booster vaccine. They were subsequently excluded from the vaccination trajectory and invited to participate in a sub-study on the immunogenicity of natural SARS-CoV-2 infection. Samples were collected 7 and 28 days after the participants tested positive. **b-d**, Detection of (ancestral) S-specific binding IgG antibodies (**b**), T-cell responses measured by IGRA (**c**), and neutralizing antibodies targeting ancestral SARS-CoV-2 and Omicron BA.1, BA.5, and XBB.1.5 variants (**d**) before, and 7 and 28 days after breakthrough infection, which was contracted before intended vaccination with the bivalent Omicron BA.5 booster vaccine (red = Ad26.COV2.S prime; blue mRNA-based prime). Data are shown in box-and-whisker plots, with the horizontal lines indicating the median, the bounds of the boxes indicating the IQR, and the whiskers indicating the range. Bold numbers above the plots represent the respective geometric mean (titer) per timepoint. The line graphs next to each panel depict a time course of the respective geometric mean values with 95% confidence intervals.

## Discussion

Here, we report that Omicron BA.1 or BA.5 bivalent booster vaccination results in rapid recall of humoral and cellular immune responses, which wane at 3 months post-vaccination. By simultaneously assessing multiple immune parameters, we found divergent immune responses after distinct COVID-19 vaccination regimens.

The immunogenicity and boosting of SARS-CoV-2-specific immune responses by Omicron BA.1 or BA.5 bivalent vaccination was in line with previous studies^7,17,19^. As published vaccination trials often do not take original priming vaccination regimens into account, no studies to our knowledge have assessed bivalent vaccine immunogenicity in the context of different priming regimens. Here, we find two important differences between bivalent-boosted participants primed with either Ad26.COV2.S or an mRNA-based vaccine: (1) mRNA-based priming leads to higher antibody levels upon boost, and (2) BA.5-bivalent boost only led to broad neutralization profiles in mRNA-primed participants. This could be related to biological differences between the vaccine platforms, as it was already shown that the vaccine effectiveness for adenovirus-vectored vaccines was lower compared to mRNA-based vaccines^3,9^

When zooming in on the booster vaccines, mRNA-1273.214 and mRNA-1273.222 proved more immunogenic than their BNT162b2 Omicron BA.1 and BA.5 counterparts. This supports a recent Moderna-funded retrospective cohort study, which reported a greater effectiveness of mRNA-1273.222 compared with BNT162b2 Omicron BA.5 in preventing COVID-19-related hospitalizations and outpatient visits^20^. The differences in immunogenicity and efficacy between the Pfizer and Moderna vaccines are likely explained by differences in dose and/or antigen design. At 3 month post-bivalent booster vaccination, we uniformly observed waning of all measured immune parameters, consistent with previous reports^21,22^. Interestingly, Omicron BA.1 and BA.5 neutralizing antibodies waned slower compared to ancestral SARS-CoV-2 neutralizing antibodies after bivalent boost. The number of antigen exposures could be underlying this observation; repeated exposure is thought to boost antibodies of the IgG4 subclass, potentially affecting functionality^23^.

Neutralizing antibodies are assumed to be the immunological correlate of protection against symptomatic SARS-CoV-2 infection^9^ and severe disease^24^. Based on this assumption, ‘variant-modified’ booster vaccinations were predicted to offer an elevated level of protection^25^. While the overall effectiveness of Omicron bivalent vaccination has been described^20,26–28^, we show that the cross-neutralization of the circulating BA.2-descendent Omicron variant XBB.1.5 was poor after administration of either the Omicron BA.1 or the BA.5 bivalent booster vaccine, in line with previous reports^4–6^. It was recently demonstrated via receptor-binding domain (RBD) depletion experiments that the immune response following an Omicron BA.5 bivalent booster vaccination is primarily ancestral-specific and only cross-reactive towards BA.5, and that the concentrations of BA.5-specific antibodies are low^29^. This is in line with a report that spike-binding monoclonal antibodies derived memory B-cells isolated from individuals boosted with variant-modified mRNA vaccines (Beta/Delta bivalent or Omicron BA.1 monovalent) predominantly recognized the ancestral SARS-CoV-2 spike protein, with only a low frequency of *de novo* B-cells targeting variant-specific epitopes^30^. Similarly, induction of new antibody responses from naïve B cells was shown to be suppressed after sequential homologous boosting^15^. As demonstrated by the low XBB.1.5 neutralizing antibody levels at 3 month post-bivalent booster independent of the prime-boost regimen, it is logical to assume that these antibodies are even less cross-reactive with potential future lineages that are antigenically even more distinct^31,32^. Their reliance on the *de novo* induction of antigen-specific B cells to maintain vaccine effectiveness may be even larger. Consequently, this argues in favor of employing monovalent vaccines based on emerging lineages in subsequent vaccination campaigns, as recently recommended by the WHO^33^.

The immunogenicity of SARS-CoV-2 breakthrough infections in comparison to booster vaccinations has not been extensively studied. Although we had a relatively small study size, and the variation between participants who had a breakthrough infection was large, we did find that breakthrough infections appear not as immunogenic as vaccination. In participants that had been enrolled but not yet vaccinated, a comparatively low boost of antibodies and rapidly waning boost of T-cell responses was detected upon breakthrough. Furthermore, in participants that had been vaccinated between 28 days and 3 months prior, no additional boost in S-specific responses was detected upon breakthrough, likely because antibody and T-cell responses were already relatively high. However, we only measured S-specific responses; breakthroughs could have potentially boosted immune responses to other antigens. Additionally, it is unknown how breakthrough infections with a variant effect protection from future infections.

Combined, our data emphasize important ‘lessons learned’ from the COVID-19 pandemic and associated vaccination strategies: (1) the original priming vaccination has an imprinting effect on the immune system that can still be observed after at least two mRNA-based booster vaccines, and (2) not all mRNA-based booster vaccines are equally immunogenic; in the SWITCH-ON trial only bivalent Omicron BA.5 vaccination broadened the neutralizing antibody response, whereas the bivalent BA.1 vaccine did not. Our data support the recent vaccination advice from the WHO (as of May 2023)^33^ to vaccinate risk groups with monovalent vaccines based on the circulating XBB.1-descendent lineage, as the current (bivalent) vaccines only induce limited cross-neutralization. Our data emphasize the importance to continuously evaluate immune responses and cross-reactivity with circulating variants to guide future COVID-19 vaccination policy making.

## Methods

### Study design and participants

The SWITCH-ON study is an ongoing multicenter, open-label, randomized controlled trial, which was conducted in accordance with the Declaration of Helsinki. Participants were randomized to either the direct boost group (DB) or the postponed boost group (PPB), who received a booster vaccination with an Omicron BA.1 or BA.5 bivalent vaccine in October or December 2022, respectively. This article reports the data for both study groups covering the period from the day of booster vaccination until 3 months post-vaccination. All participants involved in the study have given written informed consent prior to the first study visit.

HCW between the age of 18 to 65 years were invited to join the SWITCH-ON trial from four academic hospitals in the Netherlands (Amsterdam University Medical Center, Erasmus Medical Center, Leiden University Medical Center, and University Medical Center Groningen). Eligible participants were primed with either one/two dose(s) of adenovirus-based (Ad26.COV2.S) or two doses of mRNA-based vaccine (BNT162b2 or mRNA-1273), and have received at least one mRNA-based booster. Prior SARS-CoV-2 infections were allowed; however, the last booster vaccination or SARS-CoV-2 infection had to have occurred at least 12 weeks before the bivalent booster was due, as per advised interval between boosts from the National Institute for Public Health and the Environment (RIVM)^34^. Infection history was collected through a self-reported questionnaire. The full list of inclusion and exclusion criteria can be found in the study protocol^18^.

### Randomization and masking

All eligible participants were randomized using Castor software to the DB (Omicron BA.1 bivalent boost) or PPB (Omicron BA.5 bivalent boost) group in a 1:1 ratio by block randomization with block sizes of 16 and 24. Due to the set-up of the study, it was not possible to blind participants from randomization. Therefore, participants were informed about their group allocation prior to the first study visit. Randomization was completed by research assistants who were not involved in statistical analyses. Laboratory personnel were not masked from randomization allocations.

### Procedures

Participants in the DB group received an Omicron BA.1 bivalent booster in October 2022. If participants were younger than 45 years old, BNT162b2 Omicron BA.1 was administered; mRNA-1273.214 was administered to participants 45 years and older. This age division was introduced as per advice from the RIVM^34^. Following consultation with the RIVM, the age division was removed for the PPB group, and participants were randomized to receive the Omicron bivalent booster vaccination with either BNT162b2 Omicron BA.5 or mRNA-1273.222. In both groups, blood was taken during the first study visit (study visit 1, day 0). Additional blood samples were collected in subsequent study visits: study visit 2 (day 7 ± 1 days after boost), study visit 3 (day 28 ± 2 days after boost) and study visit 4 (day 90 ± 14 days after boost).

A baseline characteristics questionnaire was obtained after randomization to collect information about year of birth, biological sex, height, weight, ancestry, occupation, history of SARS-CoV-2 infection, and history of COVID-19 vaccination. A few days prior to each study visit, participants received a questionnaire to detect SARS-CoV-2 infections between the last and upcoming study visit. Via this infection questionnaire, we could identify participants who had an infection during the course of the study.

1. If the infection occurred between the informed consent session and the first vaccination study visit, participants were invited to join a sub-study to analyze immunological response after natural infection and they would be excluded from vaccination trajectory. In this sub-study, blood samples would be collected at 7 and 28 days after participants tested positive (by at-home antigen test) for COVID-19 (Figure 5).
2. If the infection occurred between baseline and day 28 post-vaccination, no additional blood samples were taken as the mixed effect of natural infection and vaccination would be difficult to distinguish. These participants were excluded from all analyses.
3. If the infection occurred between study visits day 28 and 3 months post-vaccination, participants would be invited for additional blood sampling on day 7 and 28 after they had tested positive, and remained in the study. Samples collected prior to infection were included in the immunogenicity analysis (Supplementary Figure 5).

### Outcomes

According to the study protocol, the primary outcome was the fold change (i.e., geometric mean ratio [GMR]) in antibody response between baseline and 28 days after boost across both groups. Secondary outcomes were fast response, S-specific T-cell response and levels of neutralizing antibodies^17,18^. Here, we report observational data on magnitude and quality of the immunological response. Therefore, a descriptive approach was used to describe the immunogenicity of bivalent booster vaccinations over the period of 3 months following vaccination. We measured S-specific IgG binding antibodies, S-specific T-cell responses, and neutralization of the ancestral, BA.1, BA.5, and XBB.1.5 variants. Similar parameters were analyzed in the infection sub-study.

### Identification of recent SARS-CoV-2 infection

Infections were either identified via self-reporting of participants following a positive test result in an at-home antigen test, or the detection of SARS-CoV-2 nucleocapsid (N)-specific antibodies. N-specific antibodies were measured at baseline and at 3 month post-boost using the Abbott SARS-CoV-2 IgG assay following the manufacturer’s instructions. N-specific antibody levels were expressed in a signal-to-cutoff (S/CO) ratio and the manufacturer-recommended cut-off for positivity of ≥1.4 S/CO was used. If a participants had detectable N-specific antibodies at 3 month post-boost, the other timepoints at 7 and 28 days post-boost were also tested to narrow down the moment of infection. All samples from the timepoint N-specific antibodies were detectable (or increased at least two-fold) and onwards were excluded from the immunogenicity analyses of bivalent booster vaccinations.

### Detection of SARS-CoV-2 S1-specific IgG antibodies

S1-specific antibodies were measured as previously described^35^, by Liaison SARS-CoV-2 TrimericS IgG assay (DiaSorin). The lower limit of detection (LLoD) was 4.81 BAU/mL and the cut-off for positivity was 33.8 BAU/mL, according to manufacturer’s instructions.

### Detection of SARS-CoV-2 neutralizing antibodies

Serum samples were tested for the presence of neutralizing antibodies against ancestral SARS-CoV-2, and the Omicron BA.1, BA.5, and XBB.1.5 variants in a plaque reduction neutralization test (PRNT) as previously described^17^. Viruses were cultured from clinical material and sequences were confirmed by next-generation sequencing: D614G (ancestral; GISAID: hCov-19/Netherlands/ZH-EMC-2498), Omicron BA.1 (GISAID: hCoV-19/Netherlands/LI-SQD-01032/2022), Omicron BA.5 (EVAg: 010V-04723; hCovN19/Netherlands/ZHNEMCN5892), and Omicron XBB.1.5 (GISAID: hCov-19/Netherlands/NH-EMC-5667). The human airway Calu-3 cell line (ATCC HTB-55) was used to grow virus stocks and to conduct PRNT. Calu-3 cells were cultured in OptiMEM supplemented with GlutaMAX (Gibco), penicillin (100 units/mL, Capricorn Scientific), streptomycin (0.1 mg/mL, Capricorn Scientific), and 10% fetal bovine serum (FBS; Sigma). Briefly, heat-inactivated sera were diluted two-fold serially diluted in OptiMEM without FBS. The dilutions ranges were based on the respective variant and the S-specific binding antibody level: ancestral SARS-CoV-2 (<1,500 BAU/mL: 1:10 – 1:1,280; 1,500 – 6,000 BAU/mL: 1:80 – 1:10,240; >6,000 BAU/mL: 1:640 – 1: 81,920), Omicron BA.1/BA.5 variants (<6,000 BAU/mL: 1:10 – 1:1280; >6,000 BAU/mL: 1:80 – 1: 10,240), Omicron XBB.1.5 variant (<6,000 BAU/mL: 1:10 – 1:1280; >6,000 BAU/mL: 1:40 – 1: 5,120). Four hundred PFU of either SARS-CoV-2 variant in an equal volume of OptiMEM medium were added to the diluted sera and incubated at 37°C for 1 hour. The antibody-virus mix was then transferred to Calu-3 cells and incubated at 37°C for 8 hours. Afterwards, the cells were fixed in 10% neutral-buffered formalin, permeabilized in 70% ethanol, and the plaques stained with a polyclonal rabbit anti-SARS-CoV-2 nucleocapsid antibody (Sino Biological) and a secondary peroxidase-labeled goat-anti rabbit IgG antibody (Dako). The signals were developed with a precipitate-forming TMB substrate (TrueBlue; SeraCare/KPL) and the number of plaques per well was quantified with an ImmunoSpot Image Analyzer (CTL Europe GmbH). The 50% reduction titer (PRNT50) was estimated by calculating the proportionate distance between two dilutions from which the endpoint titer was calculated. An infection control (without serum) and positive serum control (Nanogam® 100 mg/mL, Sanquin) were included on every assay plate. When no neutralization was observed, the PRNT50 was assigned a value of 10.

### Detection of T-cell responses by interferon gamma release assay (IGRA)

The SARS-CoV-2-specific T-cell response was quantified using an interferon gamma (IFN-γ) release assay (IGRA) in whole blood using the commercially available QuantiFERON SARS-CoV-2 assay kit (QIAGEN) as previously described^11^. The assay kit is certified for *in vitro* diagnostic (IVD) use. Heparinized whole blood was incubated with three different SARS-CoV-2 antigens for 20 – 24 h using a combination of peptides stimulating both CD4+ and CD8+ T-cells (Ag1, Ag2, Ag3). Mitogen- and carrier (NIL)-coated control tubes were included as positive control and negative control, respectively. After incubation, plasma was obtained by centrifugation, and IFN-γ production in response to antigen stimulation was measured by ELISA (QuantiFERON SARS-CoV-2 ELISA Kit [certified for IVD use]; QIAGEN). Results were expressed in international units (IU) IFN-γ/mL after subtraction of the NIL control values as interpolated from a standard calibration curve. LLoD was 0.01 IU/mL and the responder cut-off was 0.15 IU/mL, according to the manufacturer’s instructions. Only data obtained with Ag2 (overlapping peptides covering the ancestral S protein) is shown in this manuscript.

### Statistical analysis

A power calculation in the SWITCH-ON trial was performed to identify the number of participants required per study arm, namely: (i) Ad26.COV2.S prime in the DB group, (ii) mRNA-based prime in the DB group, (iii) Ad26.COV2.S prime in the PPB group and (iv) mRNA-based prime in the PPB group. For each arm, 91 participants were required to reach 80% power at a two-sided 5% significance level to detect a difference of 0.2 log10-transformed in the fold change of antibody response between vaccination day and 28 days after boost. This difference was based on the previous HCW study performed at Erasmus MC^35^, in which the mean fold changes for adenovirus-primed participants and mRNA-primed participants were reported as 1.344 (SD 0.451) and 1.151 (0.449), respectively.

A descriptive analysis was used to report baseline characteristics of participants. For continuous variables, mean and standard deviation (SD) were reported if the data have normal distribution. Otherwise, median and interquartile range (IQR) were used for data with non-normal distribution. Count and percentages were used to report categorical variables. For missing values, no imputation was performed and data availability was reported in **Supplementary Table S4**. Immunological data were reported as geometric mean titers or geometric means and 95% confidence intervals. Spearman correlations were reported in **Supplementary Figure S4**. No formal statistical tests were performed to test for differences within or between groups as we deviated from the original protocol in terms of pre-specified outcomes and a lower than anticipated sample size^18^.

## Data Availability

Deidentified individual participant data, the analytics code, and other supporting documents will be made available when the study is complete, upon requests made to the corresponding author.

## Acknowledgements

No private funding was received for these studies. The bivalent BA.5 vaccine mRNA-1273.222 was provided by Moderna. Moderna reviewed the final version of the manuscript, but had no role in study design, data collection, data analysis, data interpretation, or writing of the report. All other vaccines were supplied by the Center for Infectious Disease Control, National Institute for Public Health and the Environment, the Netherlands (RIVM). Cohort images were created with BioRender. This study was funded by the Netherlands Organization for Health Research and Development (ZonMw), grant agreement 10430072110001. The funder of the study had no role in study design, data collection, data analysis, data interpretation, or writing of the report.

## Author contributions statement

PHMvdK, CHGvK, and RDdV conceptualized the trial. LMZ, NHT, WJRR, DG, and RDdV performed the formal analysis. NHT, WJRR, RSGS, AG, DFP, LGV, VASHD, ML, NAK, ALWH, DvB, MPGK, PHMvdK, CHGvK, and RDdV acquired funding. All authors were involved in the investigation. LMZ, NHT, PHMvdK, CHGvK, and RDdV performed project administration. AG, DFP, LGV, PHMvdK, CHGvK, and RDdV supervised the trial. RDdV visualized the results. LMZ, NHT, PHMvdK, CGvK, and RDdV wrote the original draft of the manuscript. All authors reviewed and edited the final version of the manuscript.

## Data availability

Data from the present study are not part of public databases but are available upon reasonable request from the corresponding author. Materials and samples are available upon reasonable request, will be released via a material transfer agreement and can otherwise be obtained via

the included experimental protocols. The SARS-CoV-2 virus stocks are available through the European Virus Archive Global.

## Code availability

No specific code was written or generated for analysis of the data. Software use has been disclosed.

