## Supplementary Material for "Distinct COVID-19 vaccine combinations result in divergent immune responses"

[*] Contributed equally
[#] Contributed equally

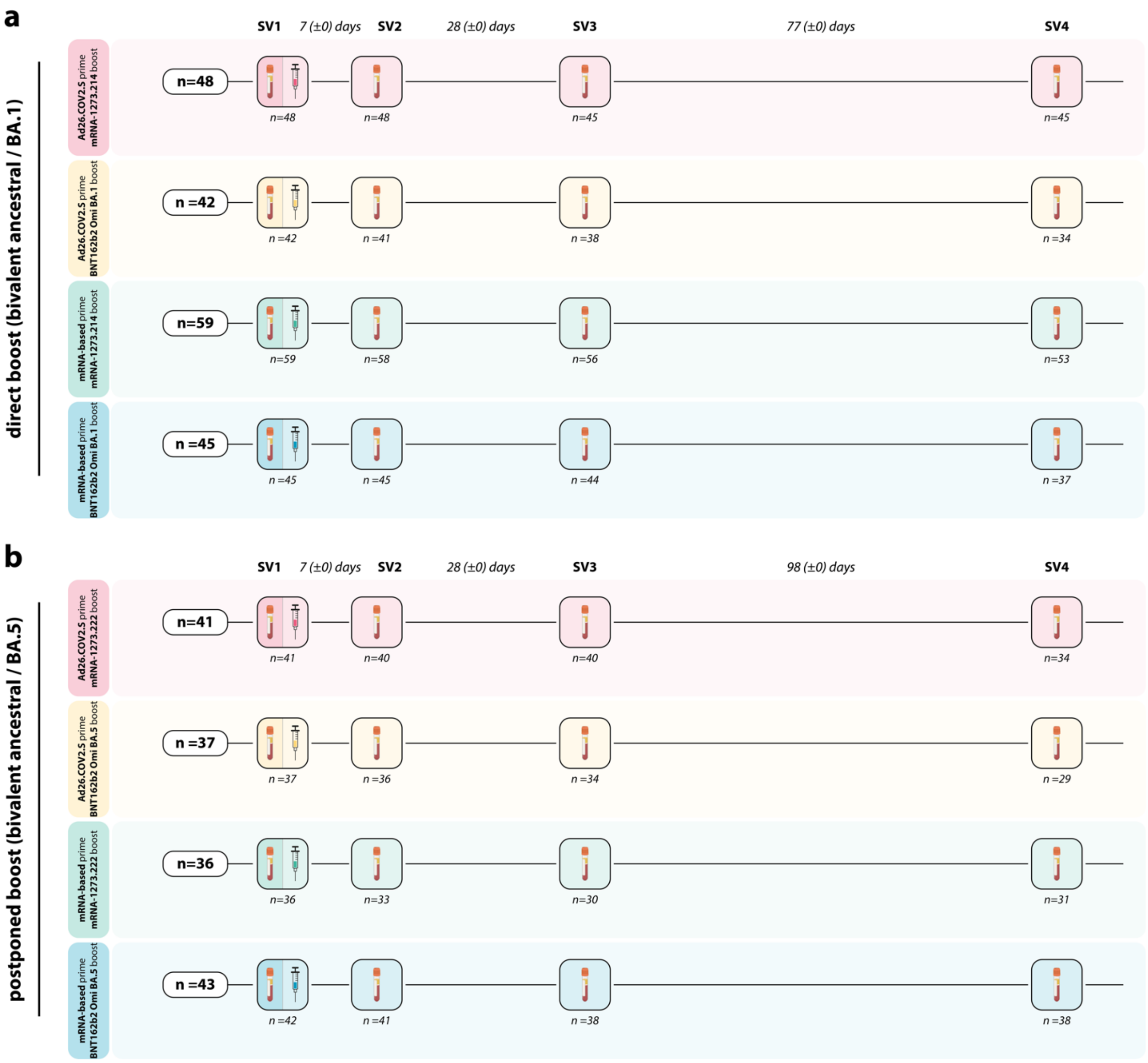


**Supplementary Figure S1. SWITCH-ON study design. a,b**, Healthcare workers (HCW), who received either Ad26.COV2.S (red and yellow) or an mRNA-based (green and blue) priming vaccination regimen, followed by an additional boost with an mRNA-based vaccine, were vaccinated with either bivalent Omicron BA.1 (**a**, direct boost group) or BA.5 (**b**, postponed boost group). Blood samples were collected at baseline before bivalent vaccination (study visit 1, SV1), at 7 (study visit 2, SV2) and 28 days post-vaccination (study visit 3, SV3), and at approximately 3 months-post vaccination (study visit 4, SV4). The numerical value in brackets indicates the IQR of days deviating from the intended interval.

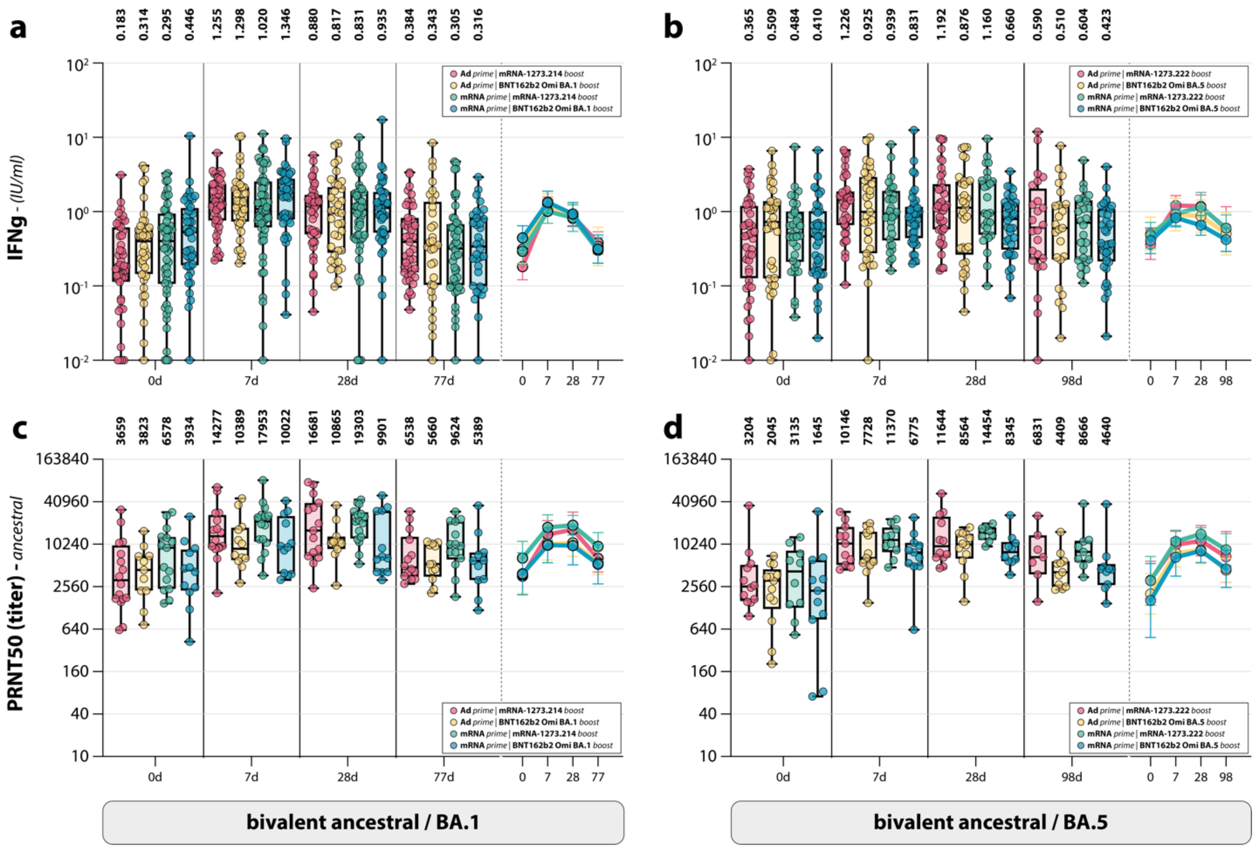


**Supplementary Figure S2. Ancestral SARS-CoV-2 neutralizing antibodies and T-cell responses in subgroups based on the different combinations of prime and boost. a-d**, Detection of T-cell responses measured by interferon-gamma (IFN-γ) release assay (IGRA) (**a**,**b**), and ancestral SARS-CoV-2 neutralizing antibodies (**c**,**d**) after Omicron BA.1 (**a**,**c**) or BA.5 (**b**,**d**) bivalent booster vaccination at baseline, and 7 days, 28 days, and 3 months post-boost. Colors indicate the specific prime-boost regimen (red = Ad26.COV2.S prime, mRNA-1273.214 or mRNA-1273.222 boost; yellow = Ad26.COV2.S prime, BNT162b2 Omicron BA.1 or BA.5 boost; green = mRNA-based prime, mRNA-1273.214 or mRNA-1273.222 boost; blue = mRNA-based prime, BNT162b2 Omicron BA.1 or BA.5 boost). Data are shown in box-and-whisker plots, with the horizontal lines indicating the median, the bounds of the boxes indicating the IQR, and the whiskers indicating the range. Bold numbers above the plots represent the respective geometric mean (titer) per timepoint. The line graphs next to each panel depict a time course of the respective geometric mean values with 95% confidence intervals.

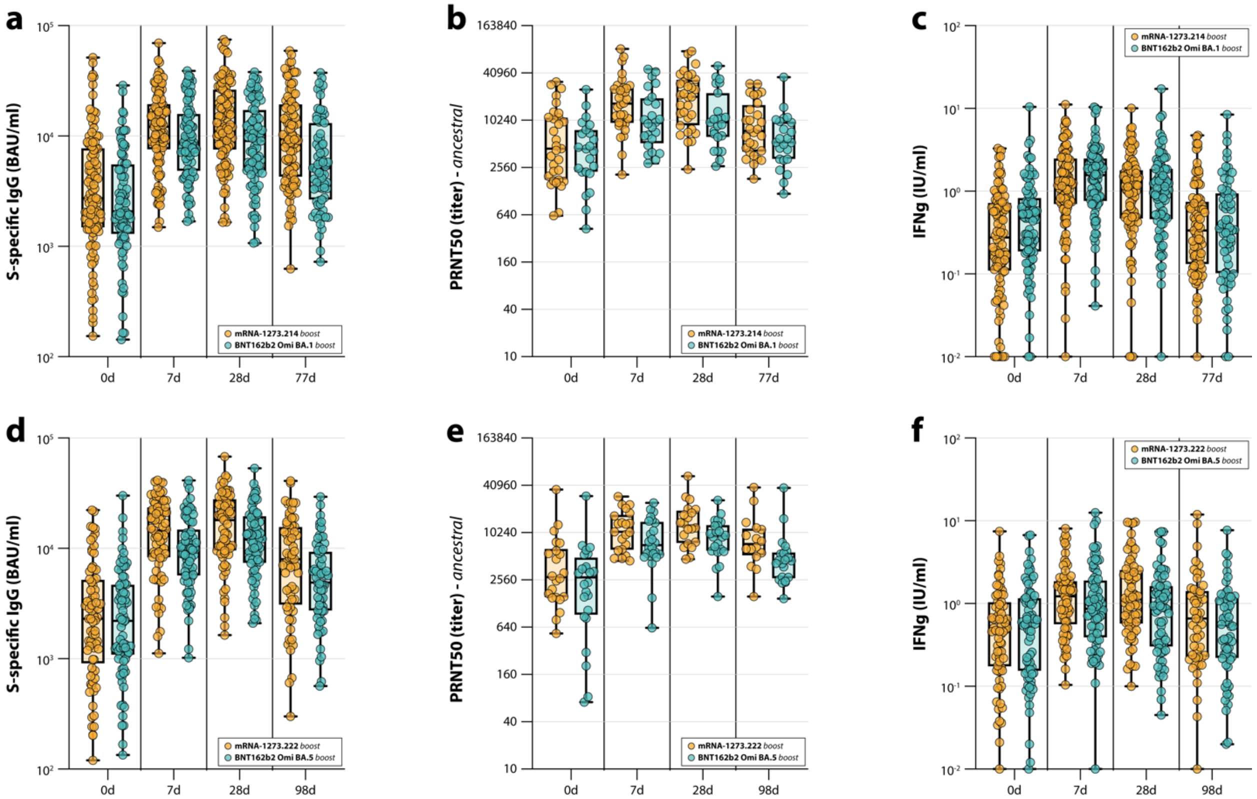


**Supplementary Figure S3. Antibody and T-cell responses after Omicron BA.5 bivalent booster vaccination separated by booster manufacturer. a-f**, Detection of (ancestral) spike (S)-specific binding IgG antibodies (**a,d**), ancestral SARS-CoV-2 neutralizing antibodies (**b,e**), and T-cell responses measured by interferon-gamma (IFN-γ) release assay (IGRA) (**c,f**) after Omicron BA.1 (**a-c**) or BA.5 (**d-f**) bivalent booster vaccination with either mRNA-1273.214 or mRNA-1273.222 (orange) or BNT162b2 Omicron BA.1 or BA.5 (teal) at baseline, and 7 days, 28 days, and 3 months post-boost. Data are shown in box-and-whisker plots, with the horizontal lines indicating the median, the bounds of the boxes indicating the IQR, and the whiskers indicating the range. Bold numbers above the plots represent the respective geometric mean (titer) per timepoint.

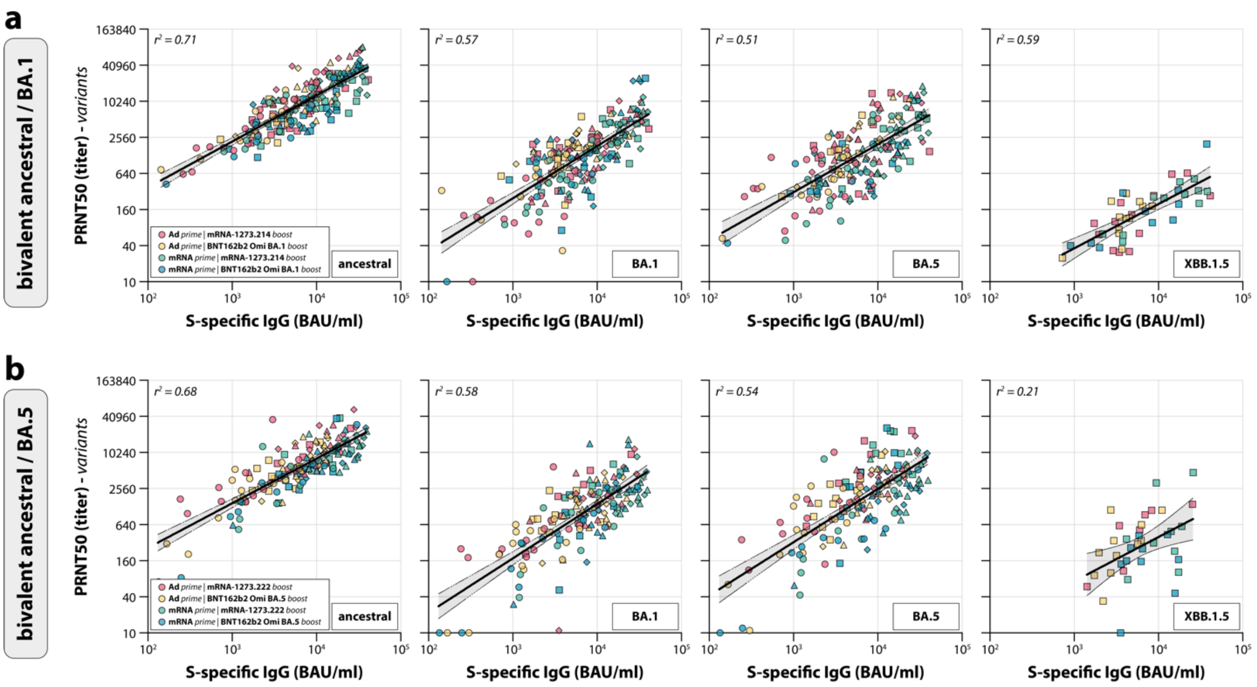


**Supplementary Figure S4. Correlations between ancestral SARS-CoV-2 and variant-specific neutralizing antibodies. a,b**, Correlations between neutralizing antibodies against ancestral SARS-CoV-2 and the Omicron BA.1, BA.5, and XBB.1.5 variants after Omicron BA.1 (**a**) or BA.5 (**b**) bivalent vaccination at 3 months post-boost. Colors indicate the specific prime-boost regimen (red = Ad26.COV2.S prime, mRNA-1273.214 or mRNA-1273.222 boost; yellow = Ad26.COV2.S prime, BNT162b2 Omicron BA.1 or BA.5 boost; green = mRNA-based prime, mRNA-1273.214 or mRNA-1273.222 boost; blue = mRNA-based prime, BNT162b2 Omicron BA.1 or BA.5 boost). Correlations were evaluated by Spearman's r.

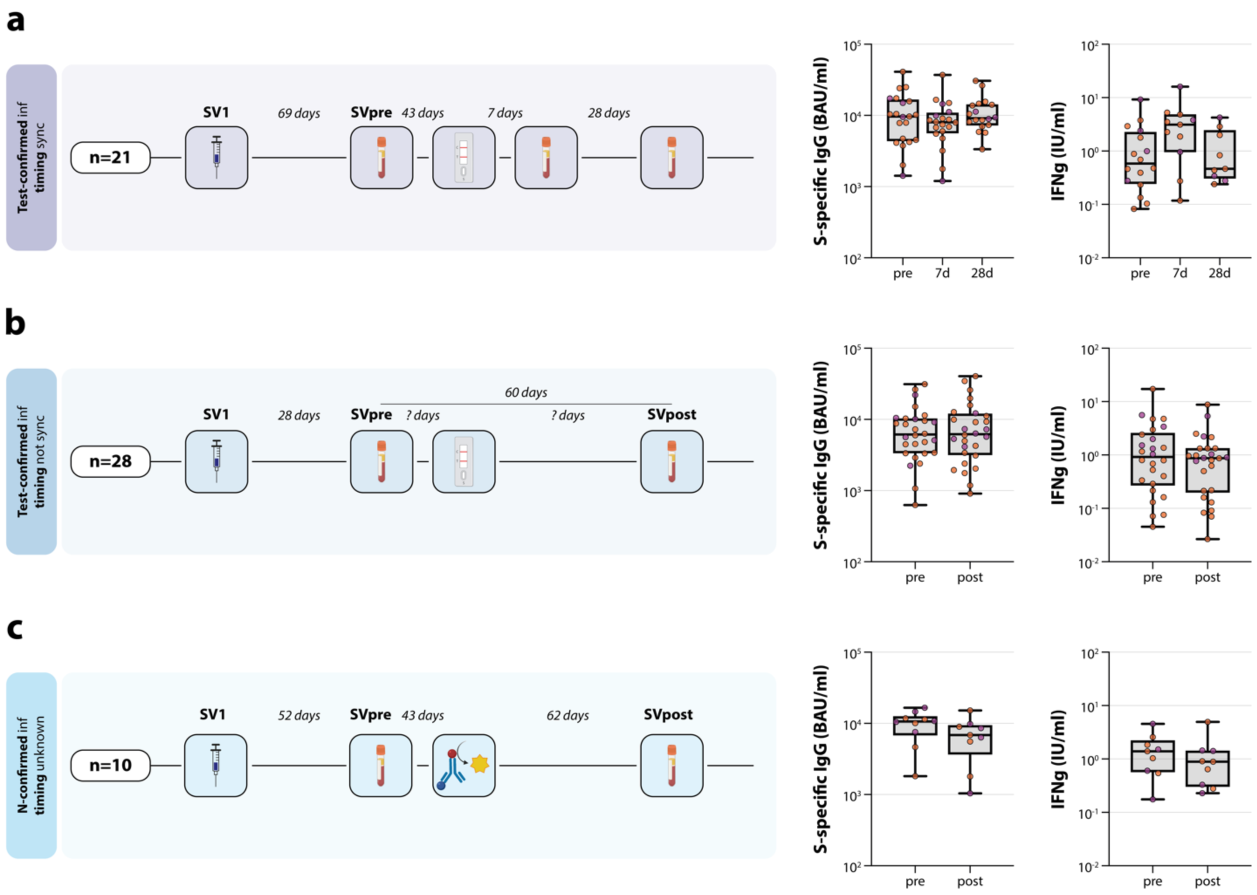


**Supplementary Figure S5. Overview of post-vaccination breakthrough infections during the SWITCH-ON trial. a-c**, Sampling procedure (left), and detection of S-specific IgG antibodies (middle) and T-cell responses measured by IGRA (right) for participants who had a breakthrough infection after Omicron BA.1 or BA.5 bivalent booster vaccination (study visit 1, SV1). Samples were either collected analogous to the pre-vaccination breakthrough infection group depicted in **Figure 5** at 7 and 28 days after testing positive (**a**), or outside of this regimen as a pre- and post-infection sample (**b,c**). Infections were either confirmed by positive test result (**a,b**), or via detection of nucleocapsid-specific antibodies (N-ELISA) (**c**).

**Supplementary Table S1.** List of investigators from the SWITCH-ON research group.

| **First name(s)** | **Surname** | **Organization** |
|---|---|---|
| Anna | van de Hoef | Erasmus MC |
| Isabelle | Veerman Roders | Erasmus MC |
| Nathalie | Tjon | Erasmus MC |
| Karenin | van Grafhorst | Erasmus MC |
| Nella | Nieuwkoop | Erasmus MC |
| Faye | de Wilt | Erasmus MC |
| Sandra | Scherbeijn | Erasmus MC |
| Babs E. | Verstrepen | Erasmus MC |
| Marion | Ferren | Erasmus MC |
| Kim | Handrejk | Erasmus MC |
| Katharina S. | Schmitz | Erasmus MC |
| Koen | Wijnans | Erasmus MC |
| Aldert C.P. | Lamoré | Erasmus MC |
| Jenny | Schnyder | Amsterdam UMC |
| Olga | Starozhitskaya | Amsterdam UMC |
| Agnes | Harskamp | Amsterdam UMC |
| Irma | Maurer | Amsterdam UMC |
| Brigitte | Boeser-Nunnink | Amsterdam UMC |
| Marga | MangasRuiz | Amsterdam UMC |
| Renate | Akkerman | UMCG |
| Martin | Beukema | UMCG |

| | | |
|---|---|---|
| Jacqueline J. | deVries-Idema | UMCG |
| Sander | Nijhof | UMCG |
| Frederique | Visscher | UMCG |
| Jopie | Zuidema | UMCG |
| Vivian W.M. | Slagter | LUMC |
| Kitty | Suijk-Benschop | LUMC |
| Jos | Fehrmann-Naumann | LUMC |
| Annelies | van Wengen-Stevenhagen | LUMC |
| Eva | Spaargaren | LUMC |
| Naomi | Olthof | LUMC |
| Anouk J.E. | de Vreede | LUMC |
| Jytte | Blokland | LUMC |

| Supplementary Table S2. Baseline characteristics of trial participants in direct boost group. | | | | | | |
|---|---|---|---|---|---|---|
| | | **Total** | **Ad26.COV2.S prime/BNT162b2 Omicron BA.1 boost** | **mRNA-based prime/BNT162b2 Omicron BA.1 boost** | **Ad26.COV2.S prime/mRNA-1273.214** | **mRNA-based prime/mRNA-1273.214** |
| | | n = 193 | n = 41 | n = 45 | n = 48 | n = 59 |
| | **Female** | 148 (77%) | 30 (73%) | 35 (78%) | 31 (65%) | 52 (88%) |
| | **Male** | 45 (23%) | 11 (27%) | 10 (22%) | 17 (35%) | 7 (12%) |
| | **Age** | 47.0 (36.0-53.0) | 36.0 (28.0-41.0) | 35.0 (28.0-40.0) | 52.5 (50.0-55.0) | 53.0 (50.0-57.0) |
| | **BMI** | 24.4 (22.6-27.4) | 23.9 (22.6-25.8) | 23.7 (21.5-26.3) | 25.2 (23.0-27.6) | 25.0 (23.1-27.7) |
| **Ancestry** | **African** | 2 (1%) | 0 (0%) | 1 (2%) | 1 (2%) | 0 (0%) |
| | **Asian** | 6 (3%) | 0 (0%) | 3 (7%) | 0 (0%) | 3 (5%) |
| | **European** | 179 (93%) | 39 (95%) | 40 (89%) | 47 (98%) | 53 (90%) |
| | **North-american** | 1 (1%) | 0 (0%) | 1 (2%) | 0 (0%) | 0 (0%) |

| | | | | | | |
|---|---|---|---|---|---|---|
| | **South-american** | 0 (0%) | 0 (0%) | 0 (0%) | 0 (0%) | 0 (0%) |
| | **Other** | 5 (3%) | 2 (5%) | 0 (0%) | 0 (0%) | 3 (5%) |
| **Occupation in hospital** | **Administrative/policy maker** | 29 (15%) | 7 (17%) | 3 (7%) | 11 (23%) | 8 (14%) |
| | **Medical doctor** | 15 (8%) | 2 (5%) | 4 (9%) | 0 (0%) | 9 (15%) |
| | **Facility services** | 3 (2%) | 1 (2%) | 1 (2%) | 1 (2%) | 0 (0%) |
| | **Management** | 24 (12%) | 4 (10%) | 3 (7%) | 9 (19%) | 8 (14%) |
| | **Supportive staff clinic/emergency department** | 2 (1%) | 1 (2%) | 0 (0%) | 0 (0%) | 1 (2%) |
| | **Supportive staff outpatient clinic** | 2 (1%) | 0 (0%) | 0 (0%) | 0 (0%) | 2 (3%) |
| | **Researcher** | 43 (22%) | 14 (34%) | 15 (33%) | 6 (13%) | 8 (14%) |
| | **Nurse** | 11 (6%) | 0 (0%) | 4 (9%) | 0 (0%) | 7 (12%) |
| | **Other** | 64 (33%) | 12 (29%) | 15 (33%) | 21 (44%) | 16 (27%) |
| **Comorbidities** | **Cardiovascular diseases** | 2 (1%) | 0 (0%) | 0 (0%) | 0 (0%) | 2 (3%) |

| | | | | | | |
|---|---|---|---|---|---|---|
| | **Pulmonary diseases** | 9 (5%) | 0 (0%) | 2 (4%) | 2 (4%) | 5 (8%) |
| | **Diabetes mellitus** | 3 (2%) | 0 (0%) | 1 (2%) | 2 (4%) | 0 (0%) |
| | **Liver diseases** | 2 (1%) | 0 (0%) | 0 (0%) | 0 (0%) | 2 (3%) |
| | **Kidney diseases** | 3 (2%) | 1 (2%) | 1 (2%) | 0 (0%) | 1 (2%) |
| **N-ELISA on study visit 1 (vaccination day)** | **Negative** | 158 (82%) | 33 (80%) | 40 (89%) | 44 (92%) | 41 (69%) |
| | **Positive** | 35 (18%) | 8 (20%) | 5 (11%) | 4 (8%) | 18 (31%) |
| **N-ELISA on study visit 4 (± 3 months post-vaccination)** | **Negative** | 150 (78%) | 30 (73%) | 36 (80%) | 42 (88%) | 42 (71%) |
| | **Positive** | 38 (20%) | 11 (27%) | 7 (16%) | 4 (8%) | 16 (27%) |
| **Original center** | **Amsterdam University Medical Center** | 21 (11%) | 5 (12%) | 8 (18%) | 3 (6%) | 5 (8%) |
| | **Erasmus Medical Center, Rotterdam** | 134 (69%) | 23 (56%) | 35 (78%) | 24 (50%) | 52 (88%) |
| | **Leiden University Medical Center** | 15 (8%) | 7 (17%) | 0 (0%) | 8 (17%) | 0 (0%) |

| | | | | | | |
|---|---|---|---|---|---|---|
| | **University Medical Center Groningen** | 23 (12%) | 6 (15%) | 2 (4%) | 13 (27%) | 2 (3%) |
| **Median time** | **Between study 1 and 2** | 7.0 (7.0-7.0) | 7.0 (7.0-7.0) | 7.0 (7.0-7.0) | 7.0 (7.0-7.0) | 7.0 (7.0-7.0) |
| | **Between study 1 and 3** | 28.0 (28.0-28.0) | 28.0 (28.0-28.0) | 28.0 (28.0-28.0) | 28.0 (28.0-28.0) | 28.0 (28.0-28.0) |
| | **Between study 1 and 4** | 77.0 (77.0-77.0) | 77.0 (77.0-77.0) | 77.0 (77.0-77.0) | 77.0 (77.0-77.0) | 77.0 (77.0-77.0) |
| | **Between last booster and bivalent booster** | 298.0 (266.0-310.0) | 267.0 (259.0-306.0) | 303.0 (297.0-310.0) | 266.0 (262.3-287.0) | 307.0 (303.0-310.0) |

*Note: categorical variables are presented as numbers (percentages), continuous variables are presented as median (interquartile range).*
*Note: the N-ELISA on study visit 4 was not performed for all participants as some did not attend this study visit. Numbers and percentages thus may slightly deviate from the total group size.*

| Supplementary Table S3. Baseline characteristics of trial participants in postponed boost group. | | | | | | |
|---|---|---|---|---|---|---|
| | | **Total** | **Ad26.COV2.S prime/BNT162b2 Omicron BA.5 boost** | **mRNA-based prime/BNT162b2 Omicron BA.5 boost** | **Ad26.COV2.S prime/mRNA-1273.222 boost** | **mRNA-based prime/mRNA-1273.222 boost** |
| | | **n = 157** | **n = 37** | **n = 43** | **n = 41** | **n = 36** |
| | **Female** | 112 (71%) | 24 (65%) | 32 (74%) | 30 (73%) | 26 (72%) |
| | **Male** | 45 (29%) | 13 (35%) | 11 (26%) | 11 (27%) | 10 (28%) |
| | **Age** | 46.0 (37.0-53.0) | 47.0 (38.0-54.0) | 48.0 (37.0-52.0) | 46.0 (36.0-54.0) | 45.0 (37.0-52.0) |
| | **BMI** | 24.6 (22.1-27.5) | 24.7 (22.1-26.9) | 24.1 (22.2-27.7) | 24.5 (22.0-25.8) | 25.5 (21.9-29.2) |
| **Ancestry** | **African** | 0 (0%) | 0 (0%) | 0 (0%) | 0 (0%) | 0 (0%) |
| | **Asian** | 4 (3%) | 0 (0%) | 1 (2%) | 1 (2%) | 2 (6%) |
| | **European** | 143 (91%) | 36 (97%) | 39 (91%) | 37 (90%) | 31 (86%) |
| | **North-american** | 0 (0%) | 0 (0%) | 0 (0%) | 0 (0%) | 0 (0%) |

| | | | | | | |
|---|---|---|---|---|---|---|
| | **South-american** | 3 (2%) | 0 (0%) | 1 (2%) | 1 (2%) | 1 (3%) |
| | **Other** | 7 (4%) | 1 (3%) | 2 (5%) | 2 (5%) | 2 (6%) |
| **Occupation in hospital** | **Administrative/policy maker** | 18 (11%) | 5 (14%) | 3 (7%) | 6 (15%) | 4 (11%) |
| | **Medical doctor** | 14 (9%) | 1 (3%) | 7 (16%) | 0 (0%) | 6 (17%) |
| | **Facility services** | 3 (2%) | 1 (3%) | 0 (0%) | 2 (5%) | 0 (0%) |
| | **Management** | 12 (8%) | 2 (5%) | 1 (2%) | 7 (17%) | 2 (6%) |
| | **Supportive staff clinic/emergency department** | 3 (2%) | 0 (0%) | 2 (5%) | 0 (0%) | 1 (3%) |
| | **Supportive staff outpatient clinic** | 1 (1%) | 0 (0%) | 1 (2%) | 0 (0%) | 0 (0%) |
| | **Researcher** | 29 (18%) | 10 (27%) | 5 (12%) | 9 (22%) | 5 (14%) |
| | **Nurse** | 13 (8%) | 1 (3%) | 8 (19%) | 0 (0%) | 4 (11%) |
| | **Other** | 64 (41%) | 17 (46%) | 16 (37%) | 17 (41%) | 14 (39%) |
| **Comorbidities** | **Cardiovascular diseases** | 4 (3%) | 0 (0%) | 1 (2%) | 1 (2%) | 2 (6%) |

| | | | | | | |
|---|---|---|---|---|---|---|
| | **Pulmonary diseases** | 7 (4%) | 4 (11%) | 1 (2%) | 0 (0%) | 2 (6%) |
| | **Diabetes mellitus** | 1 (1%) | 0 (0%) | 1 (2%) | 0 (0%) | 0 (0%) |
| | **Liver diseases** | 2 (1%) | 0 (0%) | 0 (0%) | 1 (2%) | 1 (3%) |
| | **Kidney diseases** | 0 (0%) | 0 (0%) | 0 (0%) | 0 (0%) | 0 (0%) |
| **N-ELISA on study visit 1 (vaccination day)** | **Negative** | 124 (79%) | 30 (81%) | 32 (74%) | 33 (80%) | 29 (81%) |
| | **Positive** | 32 (20%) | 7 (19%) | 10 (23%) | 8 (20%) | 7 (19%) |
| **N-ELISA on study visit 4 (± 3 months post-vaccination)** | **Negative** | 125 (80%) | 29 (78%) | 36 (84%) | 31 (76%) | 29 (81%) |
| | **Positive** | 28 (18%) | 8 (22%) | 5 (12%) | 8 (20%) | 7 (19%) |
| **Original center** | **Amsterdam University Medical Center** | 20 (13%) | 5 (14%) | 6 (14%) | 6 (15%) | 3 (8%) |
| | **Erasmus Medical Center, Rotterdam** | 107 (68%) | 21 (57%) | 36 (84%) | 19 (46%) | 31 (86%) |
| | **Leiden University Medical Center** | 13 (8%) | 6 (16%) | 0 (0%) | 7 (17%) | 0 (0%) |

| | | | | | | |
|---|---|---|---|---|---|---|
| | **University Medical Center Groningen** | 17 (11%) | 5 (14%0 | 1 (2%) | 9 (22%) | 2 (6%) |
| **Median time** | **Between study 1 and 2** | 7.0 (7.0-7.0) | 7.0 (7.0-7.0) | 7.0 (7.0-7.0) | 7.0 (7.0-7.0) | 7.0 (7.0-7.0) |
| | **Between study 1 and 3** | 28.0 (28.0-28.0) | 28.0 (28.0-28.0) | 28.0 (28.0-28.0) | 28.0 (28.0-28.0) | 28.0 (28.0-28.0) |
| | **Between study 1 and 4** | 98.0 (98.0-98.0) | 98.0 (98.0-98.0) | 98.0 (98.0-98.0) | 98.0 (98.0-98.0) | 98.0 (98.0-98.0) |
| | **Between last booster and bivalent booster** | 368.0 (336.0-483.0) | 336.0 (329.0-364.0) | 378.0 (365.5-381.0) | 337.0 (335.0-372.0) | 377.0 (372.3-381.0) |

*Note: categorical variables are presented as numbers (percentages), continuous variables are presented as median (interquartile range).*
*Note: one participant refused to disclose the their weight, therefore their BMI was excluded.*
*Note: the N-ELISA on study visit 4 was not performed for all participants as some did not attend this study visit. Numbers and percentages thus may slightly deviate from the total group size.*

**Supplementary Table S4. Data availability for both groups**

| | Direct boost group (n=193) | Postponed boost group (n=157) |
|---|---|---|
| | **Available data (%)** | **Available data (%)** |
| **Liaison - Study visit 1** | 193 (100%) | 156 (99%) |
| **Liaison - Study visit 2** | 191 (99%) | 151 (96%) |
| **Liaison - Study visit 3** | 182 (94%) | 142 (90%) |
| **Liaison - Study visit 4** | 168 (87%) | 132 (84%) |
| **Nucleocapsid - Study visit 1** | 193 (100%) | 156 (99%) |
| **Nucleocapsid - Study visit 4** | 188 (97%) | 153 (97%) |
| **IGRA Ag2 - Study visit 1** | 189 (98%) | 155 (99%) |
| **IGRA Ag2 - Study visit 2** | 190 (98%) | 146 (93%) |
| **IGRA Ag2 - Study visit 3** | 181 (94%) | 142 (90%) |
| **IGRA Ag2 - Study visit 4** | 164 (85%) | 113 (72%) |
| **PRNT50 ancestral - Study visit 1** | 54 (28%) | 46 (29%) |
| **PRNT50 ancestral - Study visit 2** | 56 (29%) | 45 (29%) |
| **PRNT50 ancestral - Study visit 3** | 54 (28%) | 42 (27%) |
| **PRNT50 ancestral - Study visit 4** | 51 (26%) | 37 (24%) |
| **PRNT50 BA.1 - Study visit 1** | 52 (27%) | 46 (29%) |
| **PRNT50 BA.1 - Study visit 2** | 53 (27%) | 43 (27%) |
| **PRNT50 BA.1 - Study visit 3** | 53 (27%) | 40 (25%) |

| | | |
|---|---|---|
| **PRNT50 BA.1 - Study visit 4** | 50 (26%) | 37 (24%) |
| **PRNT50 BA.5 - Study visit 1** | 51 (26%) | 46 (29%) |
| **PRNT50 BA.5 - Study visit 2** | 51 (26%) | 42 (27%) |
| **PRNT50 BA.5 - Study visit 3** | 48 (25%) | 39 (25%) |
| **PRNT50 BA.5 - Study visit 4** | 46 (24%) | 37 (24%) |
| **PRNT50 XBB.1.5 - Study visit 1** | 0 (0%) | 0 (0%) |
| **PRNT50 XBB.1.5 - Study visit 2** | 0 (0%) | 0 (0%) |
| **PRNT50 XBB.1.5 - Study visit 3** | 0 (0%) | 0 (0%) |
| **PRNT50 XBB.1.5 - Study visit 4** | 48 (25%) | 37 (24%) |
